# Machine learning model for predicting left atrial thrombus or spontaneous echo contrast in non-valvular atrial fibrillation patients based on multimodal echocardiographic parameters

**DOI:** 10.1101/2024.04.10.24305639

**Authors:** Decai Zeng, Shuai Chang, Xiaofeng Zhang, Xiangling Cao, Yanfen Zhong, Yongzhi Cai, Tongtong Huang, Ji Wu

**Author notes:** **Correspondence to:** Ji Wu, MD,PhD, Department of Ultrasonic Medicine, the First Affiliated Hospital of Guangxi Medical University 6 Shuangyong Road, Nanning, Guangxi Zhuang Autonomous Region, China.

## Abstract

**BACKGROUND:** Although clinical prediction models have been proposed to predict thrombosis risk in non-valvular atrial fibrillation (NVAF), machine learning (ML) based models to predict thrombotic risk were limited. This study aimed to develop a robust ML-based predictive model that integrates multimodal echocardiographic data and clinical risk factors to evaluate the risk of thrombosis in patients with NVAF.

**METHODS AND RESULTS:** A total of 402 NVAF patients scheduled for AF radiofrequency ablation and/or left atrial appendage closure at the First Affiliated Hospital of Guangxi Medical University from January 2020 to December 2023 were prospectively collected. Among them, there were 289 males (71.9%) and 113 females (28.1%), with a mean age of 59.7 years. There were 142 patients (35.3%) with left atrial thrombus/spontaneous echocardiographic contrast (LAT/SEC) and 260 patients (64.7%) without LAT/SEC. Clinical data, biochemical markers, and multimodal echocardiographic parameters were collected to construct the model. After screening the influencing factors with Least Absolute Shrinkage and Selection Operator (LASSO) regression, we explored seven ML models – Logistic Regression (LR), Decision Tree (DT), K-Nearest Neighbors (KNN), Light Gradient Boosting Machine (LightGBM), Support Vector Machine (SVM), Random Forest (RF), and eXtreme Gradient Boosting (XGBoost), to forecast the risk of thrombosis in NVAF patients. A variety of metrics, such as accuracy, precision, recall, F1 score and area under the curve (AUC) were used to evaluate the performance of the models. The DeLong test was applied to compare area under receiver operating characteristic (AUROC) curves across different ML models. Decision curve analysis (DCA) was used to gauge the clinical utility of each ML model by comparing their clinical net benefit. To gain insight into the risk prediction model, we used Shapley additive explanations (SHAP) analysis and investigated the contributions of the different variables. The incorporation of multimodal echocardiographic parameters and clinical risk factors using advanced ML algorithms markedly enhanced the accuracy of predicting thrombosis risk in individuals with NVAF. Specifically, the XGBoost model (AUC 0.959, 95% CI 0.925–0.993) slightly outperformed the traditional LR model (AUC 0.949, 95% CI: 0.911-0.987) in predicting thrombosis risk in NVAF patients, and showed superior predictive ability compared to other ML algorithms. Additionally, XGBoost offered greater clinical net benefit within a threshold probability range of 0.1 to 1.0. SHAP analysis revealed that left atrial structure (left atrial volume index, three-dimensional sphericity index), hemodynamic parameters (left atrial acceleration factor and S/D ratio), and functional parameters (peak atrial longitudinal strain and left ventricular ejection fraction) were important features in predicting the risk of thrombus formation in NVAF patients, with reduced peak atrial longitudinal strain being the most important risk factor for predicting thrombus.

**CONCLUSIONS:** Developing a predictive model utilizing ML techniques that incorporate multimodal echocardiographic parameters in conjunction with clinical risk factors has the potential to enhance the predictive accuracy of the thrombosis risk in individuals with NVAF. The XGBoost model shows that decreased PALS, hemodynamic abnormalities and left atrium spherical remodeling are significant factors correlated with increased risk of thrombus in NVAF.

## Introduction

Atrial fibrillation (AF) is a common clinical arrhythmia, with stroke being the predominant thrombotic complication in non-valvular atrial fibrillation (NVAF) and a major contributor to disability.^1^ The dislodgement of thrombi from the left atrium or left atrial appendage is the primary etiology of stroke in NVAF patients. Thus, a precise evaluation of thrombosis risk in the left atrium and left atrial appendage is crucial for effective stroke prevention in NVAF patients.^2^ This risk evaluation not only guides clinical management decisions but also has the potential to significantly improve patient outcomes

Clinical practice frequently suggests utilizing the CHA_2_DS_2_-VASc score for assessing stroke risk, which primarily relies on clinical risk factors and does not incorporate biochemical markers.^3^ In contrast, the ABC score incorporates biochemical markers such as NT-proBNP and troponin I, and is a novel scoring system developed from extensive cohort studies. It is user-friendly and offers enhanced risk assessment capabilities when compared to the commonly utilized CHA₂DS₂-VASc score.^4^ This advancement holds significant implications for clinical diagnosis, evaluation, and medication management. Nevertheless, this score fails to account for indicators such as left atrial morphology and functional parameters, thus presenting certain limitations.

Changes in left atrial structure, function, and hemodynamics are instrumental for AF prediction and stroke risk evaluation.^5^ A detailed assessment of left atrial function using echocardiography has been shown to be beneficial in assessing risk, notably in NVAF patients with low CHA_2_DS_2_-VASc scores.^6^ Additionally, left atrial strain serves as a valuable surrogate indicator of structural remodeling and fibrosis.^7^ Studies suggest that combining echocardiographic measurement of left atrial strain with the CHA_2_DS_2_-VASc score can provide additional insight into stroke risk, helping with decisions about anticoagulation in newly diagnosed NVAF patients.

Machine learning (ML), with its novel applications in medicine, has demonstrated significant promise in cardiovascular disease assessment.^8,9^ The rise of machine learning technologies, especially when combined with multi-modal echocardiographic parameters, provides new perspectives for clinical risk assessment.^10^ Prediction models of thrombosis risk in atrial fibrillation (AF) are used to guide treatment.^11^ Although regression models have traditionally been the preferred analytical approach for prediction modeling, ML has emerged as a potentially more effective methodology. In this study, we aim to develop a predictive model that employs machine learning (ML) techniques in conjunction with detailed echocardiographic parameters and essential clinical factors. And compare the predictive accuracy of this ML-based model with that of traditional regression models in determining the risk of thrombosis among patients with NVAF.

## Methods

### Study population

We performed a prospective, continuous study collecting data from inpatients at the First Affiliated Hospital of Guangxi Medical University. The study was conducted from January 2020 to December 2023 and focused on patients who were scheduled to undergo AF radiofrequency ablation and/or left atrial appendage closure. The objective was to develop a machine learning model to predict left atrial thrombus (LAT) or spontaneous echo contrast (SEC) in patients with NVAF. Inclusion Criteria: Patients diagnosed with paroxysmal or persistent AF; Aged 18 years or older; Capable of undergoing transesophageal echocardiography. Exclusion Criteria: Presence of congenital heart disease; Myocardial disease; heart valve disease; History of valve repair or replacement surgery; Acute coronary syndrome; Moderate to severe functional mitral regurgitation. The research protocol has received approval from the Ethics Committee of the First Affiliated Hospital of Guangxi Medical University (Approval No: 2022-KT-077). All NVAF patients involved in the study have provided signed informed consent. The flowchart of the study participants and Schematic of study design was shown in Figure 1.

**Figure 1.**
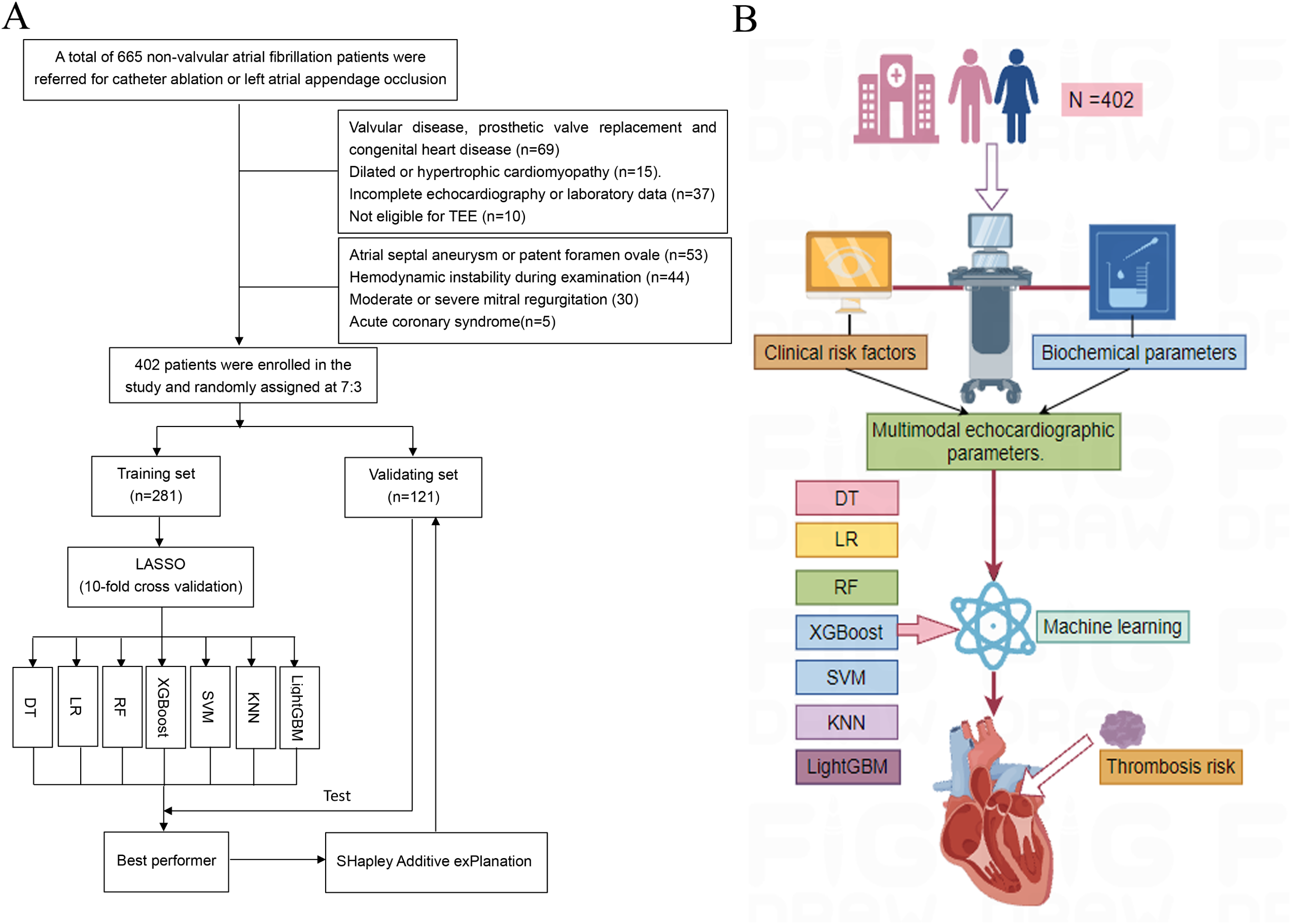
(A) Flowchart of the participants’ selection and study design. (B) Graphical abstract (DT, Decision Tree; KNN, K-Nearest Neighbors; LightGBM, Light Gradient Boosting Machine; LR, Logistic Regression; NVAF, non-valvular atrial fibrillation; RF, random forest; RF, Random Forest; TEE, transesophageal echocardiography; SVM, Support Vector Machine; XGBoost, eXtreme Gradient Boosting).

### Risk Factor Assessment and Laboratory Examination

Access the electronic medical records system to gather basic patient information including age, gender, body surface area, blood pressure, and more. Past medical history should include conditions like hypertension, coronary heart disease, diabetes, hyperlipidemia, peripheral vascular disease, history of stroke or transient ischemic attack (TIA), heart failure, and AF type. Calculate the CHA_2_DS_2_-VASc score based on clinical history and other information. Also record history of anticoagulant drug use, including antiplatelet agents, warfarin, and new oral anticoagulants. After admission, fasting venous blood samples are taken and analyzed by the hospital laboratory to obtain biochemical indicators such as N-terminal pro-brain natriuretic peptide (NT-proBNP), cardiac troponin I, serum creatinine (SCr), endogenous creatinine clearance rate (Ccr), and more. Calculate the glomerular filtration rate (eGFR) using the CKD-EPI formula^12^, which considers factors like serum creatinine (SCr) levels, age, gender, and race. In total, 22 clinical data and biochemical indicators are collected.

### Echocardiography

Transthoracic echocardiography (TTE) and transesophageal echocardiography (TEE) were carried out using the Philips EPIQ 7C echocardiographic diagnostic system from Koninklijke Philips N.V. in the Netherlands. TTE utilized the S5-1 probe (1 to 5 MHz), while TEE utilized the X8-2t probe (2 to 8 MHz). We ensured that the time between TTE and TEE assessments did not exceed 48 hours to maintain consistency and reliability in the cardiac evaluation.

Measurements of the cardiac chambers’ linear dimensions followed standard methods.^13^ The left atrial diameter is measured in the parasternal long axis view of the left ventricle. Additionally, the left atrial transverse and longitudinal diameters are captured in the apical four-chamber view. We calculate the 2D sphericity index (2D-SI) of the left atrium using the ratio of its transverse diameter to the longitudinal diameter. The left ventricular mass was determined using the Devereux formula: LVM = 0.8 {1.04[(SWT+LVEDD+PWT)^3^−LVEDD^3^]}+0.6, where SWT represents the septal wall thickness at end-diastole, LVEDD the left ventricular end-diastolic diameter, and PWT the posterior wall thickness at end-diastole.

Furthermore, a transthoracic three-dimensional (3D) matrix transducer is employed to acquire detailed images of the left heart in the standard apical four-chamber view. The intelligent Heart Model (HM) functionality was used to gather 3D volumetric data, which includes the left ventricular end-diastolic volume (LVEDV), end-systolic volume (LVESV), ejection fraction (EF), and the left atrial volume (LAV). The left atrial maximum diameter (LADmax) is determined by identifying the largest measurement among the internal, transverse, and longitudinal diameters of the left atrium. The three-dimensional sphericity index (3D-SI) is then calculated by comparing the LAV to the volume of a sphere that has LADmax as its diameter (Figure 2A-C). In order to adjust for differences in body size, the left atrial volume and left ventricular mass were normalized to the body surface area (BSA), resulting in the left atrial volume index (LAVI) and left ventricular mass index (LVMI).

**Figure 2.**
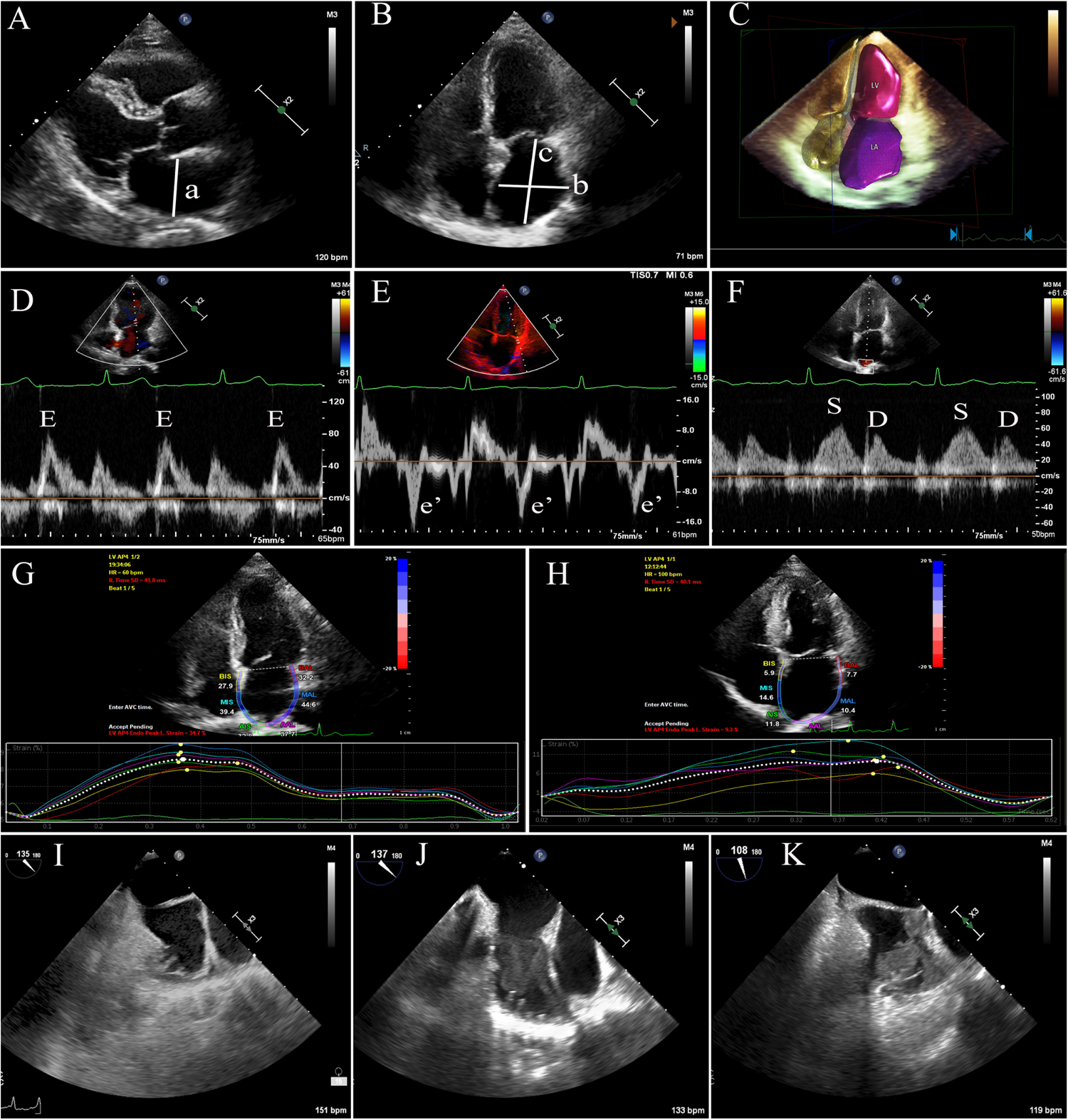
Demonstration of measuring multiple parameters using echocardiography and recognition of left atrial thrombus/spontaneous contrast. (A-C), Measurements of left atrial diameter (a), transverse diameter(b), longitudinal diameter(c), and left atrial three-dimensional volume. (D), Mitral valve peak early diastolic E-wave flow velocity. (E), Mitral annular lateral wall early diastolic velocity (e’). (F) The pulmonary venous flow velocity spectrum shows that the S-wave represents the systolic peak blood flow velocity and the D-wave represents the diastolic peak blood flow velocity. (G-H) Normal left atrial peak longitudinal strain and decreased left atrial peak longitudinal strain. (I), without left atrial thrombus or spontaneous echo contrast, (J), spontaneous echo contrast, (K) left atrial thrombus.

We performed left atrial strain measurements using two-dimensional speckle tracking technology. Standard dynamic images were acquired from the apical four-chamber view and saved in the Dicom format for subsequent analysis. Off-line analysis was conducted using QLAB software, where the measurement process begins with tracing the endocardial border of the left atrium. Starting from one side of the mitral annulus, the trace proceeds along the inner border, avoiding pulmonary veins and the left atrial appendage, and ends at the opposing side of the mitral annulus. The region of interest (ROI) was set at a width of 3 mm. Adjustments to the ROI’s size and shape were made to encompass the full thickness of the left atrial wall, deliberately excluding the pericardium. From this, we obtained the left atrial strain curve and specifically measured the peak atrial longitudinal strain (PALS)

We recorded the peak early diastolic mitral flow velocity (E) in the apical four-chamber view and calculated the E/e’ ratio. Additionally, we determined the S/D ratio using the S and D velocities from the left atrial inflow at the right upper pulmonary vein. In AF rhythm, echocardiographic measurements—such as E peak, e’ velocity, and pulmonary vein flow spectra—were averaged over five cardiac cycles for consistency. For patients in sinus rhythm, Doppler measurements were averaged over three cycles. The left atrial acceleration factor (α) was defined as α = E/[(S+D)/2], with E representing the peak E flow at the mitral valve, and S and D denoting peak flow velocities at the pulmonary vein.^14^

Before TEE, patients were educated on the procedure’s purpose and process and provided informed consent. A left atrial/left atrial appendage thrombus (LAT) was identified as a low-echoic mass, distinct from the endocardium and trabeculae, and observable across multiple TEE views. Additionally, SEC—smoke-like shadows within the left atrium or appendage—was evaluated after gain optimization as a precursor to thrombus formation, indicative of stagnant flow and a hypercoagulable state. Demonstration of measuring multiple parameters using echocardiography and recognition of left atrial thrombus/spontaneous contrast were shown in Figure 2.

All echocardiographic parameters were taken by expert cardiac physician, and to ensure objectivity, those involved in measuring echocardiographic indices and analyzing data were kept blinded to patient details.

### Prediction model

This study divided the NVAF patients into two groups: a training set with 70% of the sample and a test set with the remaining 30%. We used Least Absolute Shrinkage and Selection Operator (LASSO) regression for feature selection and conducted 10-fold cross-validation to optimize the training of the classification algorithm on the training set, identifying the best subset of variables for the predictive model. To evaluate feature importance, the study analyzed the intrinsic logic of the ML models, with feature weights indicating their impact on prediction. Feature importance charts visually depicted these findings, enhancing transparency and interpretability.

We explored seven machine learning (ML) models – Logistic Regression (LR), Decision Tree (DT), K-Nearest Neighbors (KNN), Light Gradient Boosting Machine (LightGBM), Support Vector Machine (SVM), Random Forest (RF), and eXtreme Gradient Boosting (XGBoost) – to forecast the risk of thrombosis in NVAF patients.^15,16^ An array of metrics, including accuracy, precision, recall, F1 score, area under the curve (AUC), and 95% confidence intervals (CI), were employed to assess the models’ performance. ‘Accuracy’ measures the model’s ability to correctly identify cases (true positives and true negatives). ‘Precision’ assesses the ratio of correctly predicted positive cases to all predicted positive cases, while ‘recall’ evaluates the model’s capability to detect true positives. The ‘F1 score’ acts as a combined metric of precision and recall, offering a comprehensive evaluation of the model’s performance.

Furthermore, SHAP (SHapley Additive exPlanation) analysis was utilized to elucidate the predictive efficacy and influence of specific features on the model’s results, drawing upon game theory principles.^17^ SHAP delves into the rationale behind each prediction, facilitating a comprehensive comprehension of feature impacts and revealing their positive or negative associations with predicted outcomes. The visual aids provided by SHAP offer lucid depictions of feature contributions for individual instances, as well as their broader significance within the dataset.

### Statistical analyses

Continuous variables were first assessed for normality using the Shapiro-Wilk test. Variables following a normal distribution were described using as mean and standard deviation (SD) and compared via Student’s t-tests. Non-normally distributed variables were summarized using medians and interquartile ranges, with the Mann-Whitney test employed for comparisons between groups. Categorical variables were presented as frequencies and percentages, and analyzed using either Pearson’s chi-square test or Fisher’s exact test, as appropriate. For machine learning (ML) analyses, we utilized the R programming language. Specific ML models were implemented using various R packages: LASSO regression with the “glmnet” package, decision trees with the “rpart” package, random forests with the “randomForest” package, XGBoost with the “xgboost” package, support vector machines (SVM) with the “e1071” package, k-nearest neighbors (knn) with the “kknn” package, and LightGBM with the “lightgbm” package. The DeLong test was applied to compare receiver operating characteristic (ROC) curves across different ML models. Decision curve analysis (DCA) was used to gauge the clinical utility of each ML model by comparing their clinical net benefit. Statistical analyses were carried out using SPSS version 25.0 and R statistical language (Version R 4.2.1). A P-value less than 0.05 was considered as indicative of statistical significance.

## Results

This study included a total of 402 patients with NVAF who met the inclusion and exclusion criteria, all of whom participated in the construction and validation phases of a thrombosis risk prediction model. The cohort consisted of 289 males (71.9%) and 113 females (28.1%), with an average age of 59.7 ± 10.8 years. In our study, 142 patients presented with LAT/SEC (35.3%), while 260 patients were absence of LAT/SEC (64.7%).

A range of 34 characteristic parameters were assessed in this study, encompassing general clinical information—17 variables in total—including age, gender, BSA, systolic and diastolic blood pressure, presence of hypertension, diabetes, hyperlipidemia, history of stroke/TIA, CAD, heart failure, type of AF, CHA_2_DS_2_-VASc score, and a history of anticoagulant or antiplatelet drug use (antiplatelet drugs, warfarin, and novel oral anticoagulants). Biochemical measurements were comprised of five variables: Scr, Ccr, eGFR, NT-proBNP and troponin I. In terms of multimodal echocardiographic parameters, 12 variables were analyzed including LVEDV, LVESV, LVEF, LVMI, LAVI, 2D-SI, 3D-SI, E, E/e’ ratio, S/D ratio, left atrial acceleration factor (α), and PALS etc.

The comparative analysis of these parameters between patients with and without LAT/SEC was presented in Table 1. In this research, 34 characteristic variables were initially considered, with LASSO regression employed for variable selection. The LASSO regression method applied a penalty function that shrinks some coefficients towards zero, effectively compressing them while ensuring the sum of their absolute values remains below a predetermined threshold. This process resulted in some coefficients being set to zero, thereby enabling the generation of a simplified and more interpretable model. Utilizing ten-fold cross-validation, the study automatically excluded variables that had zero-valued coefficients through this method. Following this process, 9 key variables were selected, which included persistent AF, history of stroke/TIA, Ccr, EF, LAVI, 3D-SI, pulmonary vein S/D ratio, left atrial acceleration factor (α), and PALS. The finalized set of predictive variables and their selection process were illustrated in Figure 3.

**Table 1.**
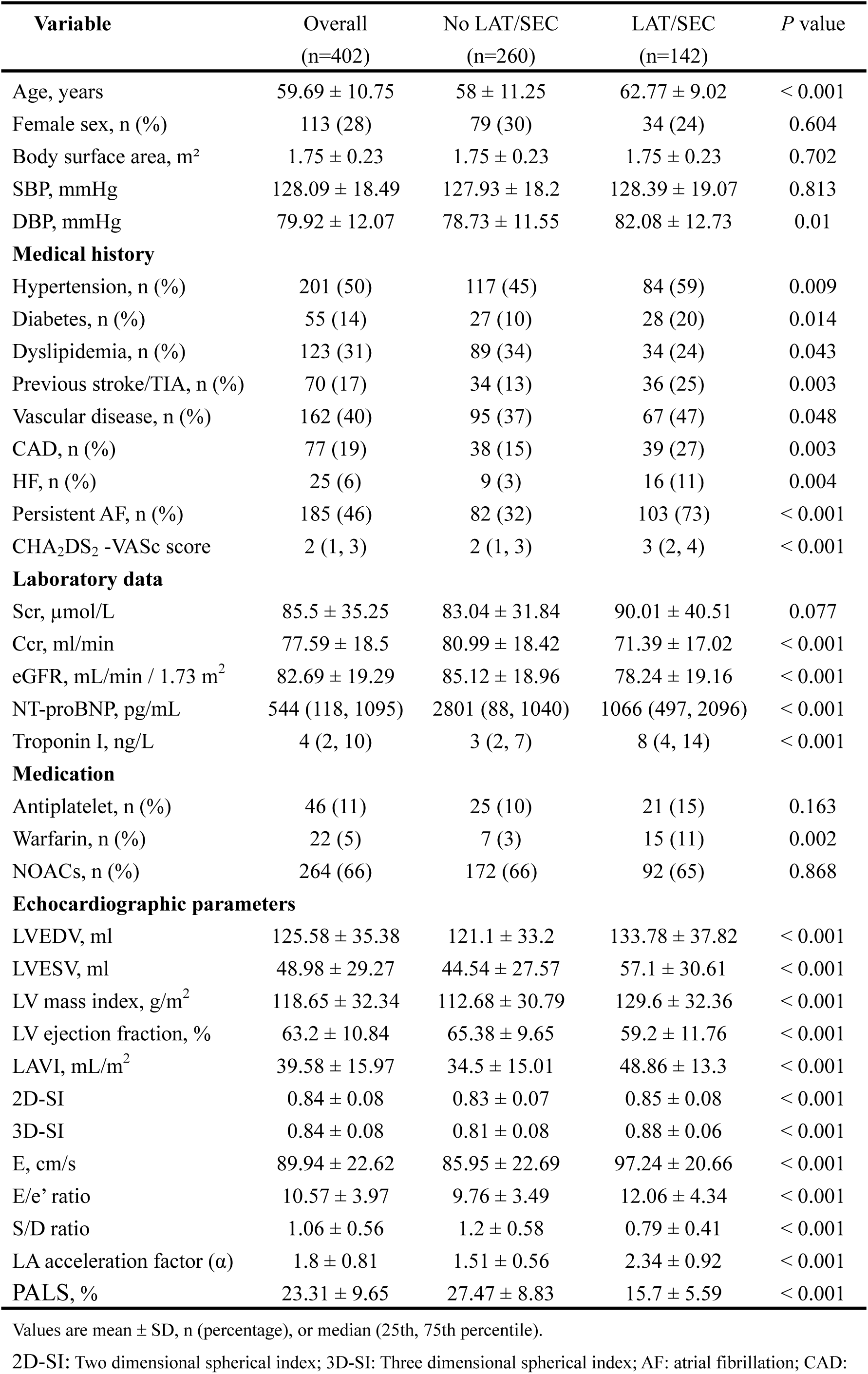
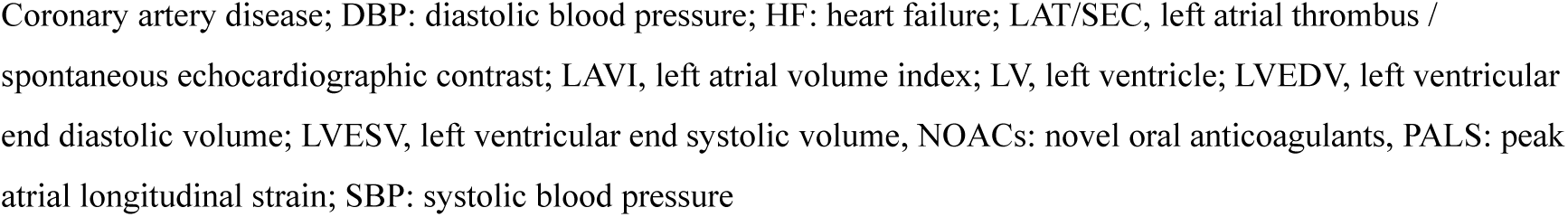
Baseline Characteristics of the 402 patients stratified by LAT/SEC.

| Variable | Overall<br>(n=402) | No LAT/SEC<br>(n=260) | LAT/SEC<br>(n=142) | P value |
| --- | --- | --- | --- | --- |
| Age, years | 59.69 ± 10.75 | 58 ± 11.25 | 62.77 ± 9.02 | < 0.001 |
| Female sex, n (%) | 113 (28) | 79 (30) | 34 (24) | 0.604 |
| Body surface area, m<sup>2</sup> | 1.75 ± 0.23 | 1.75 ± 0.23 | 1.75 ± 0.23 | 0.702 |
| SBP, mmHg | 128.09 ± 18.49 | 127.93 ± 18.2 | 128.39 ± 19.07 | 0.813 |
| DBP, mmHg | 79.92 ± 12.07 | 78.73 ± 11.55 | 82.08 ± 12.73 | 0.01 |
| **Medical history** |  |  |  |  |
| Hypertension, n (%) | 201 (50) | 117 (45) | 84 (59) | 0.009 |
| Diabetes, n (%) | 55 (14) | 27 (10) | 28 (20) | 0.014 |
| Dyslipidemia, n (%) | 123 (31) | 89 (34) | 34 (24) | 0.043 |
| Previous stroke/TIA, n (%) | 70 (17) | 34 (13) | 36 (25) | 0.003 |
| Vascular disease, n (%) | 162 (40) | 95 (37) | 67 (47) | 0.048 |
| CAD, n (%) | 77 (19) | 38 (15) | 39 (27) | 0.003 |
| HF, n (%) | 25 (6) | 9 (3) | 16 (11) | 0.004 |
| Persistent AF, n (%) | 185 (46) | 82 (32) | 103 (73) | < 0.001 |
| CHA<sub>2</sub>DS<sub>2</sub> -VASc score | 2 (1, 3) | 2 (1, 3) | 3 (2, 4) | < 0.001 |
| **Laboratory data** |  |  |  |  |
| Scr, µmol/L | 85.5 ± 35.25 | 83.04 ± 31.84 | 90.01 ± 40.51 | 0.077 |
| Ccr, ml/min | 77.59 ± 18.5 | 80.99 ± 18.42 | 71.39 ± 17.02 | < 0.001 |
| eGFR, mL/min / 1.73 m<sup>2</sup> | 82.69 ± 19.29 | 85.12 ± 18.96 | 78.24 ± 19.16 | < 0.001 |
| NT-proBNP, pg/mL | 544 (118, 1095) | 2801 (88, 1040) | 1066 (497, 2096) | < 0.001 |
| Troponin I, ng/L | 4 (2, 10) | 3 (2, 7) | 8 (4, 14) | < 0.001 |
| **Medication** |  |  |  |  |
| Antiplatelet, n (%) | 46 (11) | 25 (10) | 21 (15) | 0.163 |
| Warfarin, n (%) | 22 (5) | 7 (3) | 15 (11) | 0.002 |
| NOACs, n (%) | 264 (66) | 172 (66) | 92 (65) | 0.868 |
| **Echocardiographic parameters** |  |  |  |  |
| LVEDV, ml | 125.58 ± 35.38 | 121.1 ± 33.2 | 133.78 ± 37.82 | < 0.001 |
| LVESV, ml | 48.98 ± 29.27 | 44.54 ± 27.57 | 57.1 ± 30.61 | < 0.001 |
| LV mass index, g/m<sup>2</sup> | 118.65 ± 32.34 | 112.68 ± 30.79 | 129.6 ± 32.36 | < 0.001 |
| LV ejection fraction, % | 63.2 ± 10.84 | 65.38 ± 9.65 | 59.2 ± 11.76 | < 0.001 |
| LAVI, mL/m<sup>2</sup> | 39.58 ± 15.97 | 34.5 ± 15.01 | 48.86 ± 13.3 | < 0.001 |
| 2D-SI | 0.84 ± 0.08 | 0.83 ± 0.07 | 0.85 ± 0.08 | < 0.001 |
| 3D-SI | 0.84 ± 0.08 | 0.81 ± 0.08 | 0.88 ± 0.06 | < 0.001 |
| E, cm/s | 89.94 ± 22.62 | 85.95 ± 22.69 | 97.24 ± 20.66 | < 0.001 |
| E/e' ratio | 10.57 ± 3.97 | 9.76 ± 3.49 | 12.06 ± 4.34 | < 0.001 |
| S/D ratio | 1.06 ± 0.56 | 1.2 ± 0.58 | 0.79 ± 0.41 | < 0.001 |
| LA acceleration factor (α) | 1.8 ± 0.81 | 1.51 ± 0.56 | 2.34 ± 0.92 | < 0.001 |
| PALS, % | 23.31 ± 9.65 | 27.47 ± 8.83 | 15.7 ± 5.59 | < 0.001 |

Values are mean ± SD, n (percentage), or median (25th, 75th percentile).
2D-SI: Two dimensional spherical index; 3D-SI: Three dimensional spherical index; AF: atrial fibrillation; CAD:
Coronary artery disease; DBP: diastolic blood pressure; HF: heart failure; LAT/SEC, left atrial thrombus / spontaneous echocardiographic contrast; LAVI, left atrial volume index; LV, left ventricle; LVEDV, left ventricular end diastolic volume; LVESV, left ventricular end systolic volume, NOACs: novel oral anticoagulants, PALS: peak atrial longitudinal strain; SBP: systolic blood pressure

**Figure 3.**
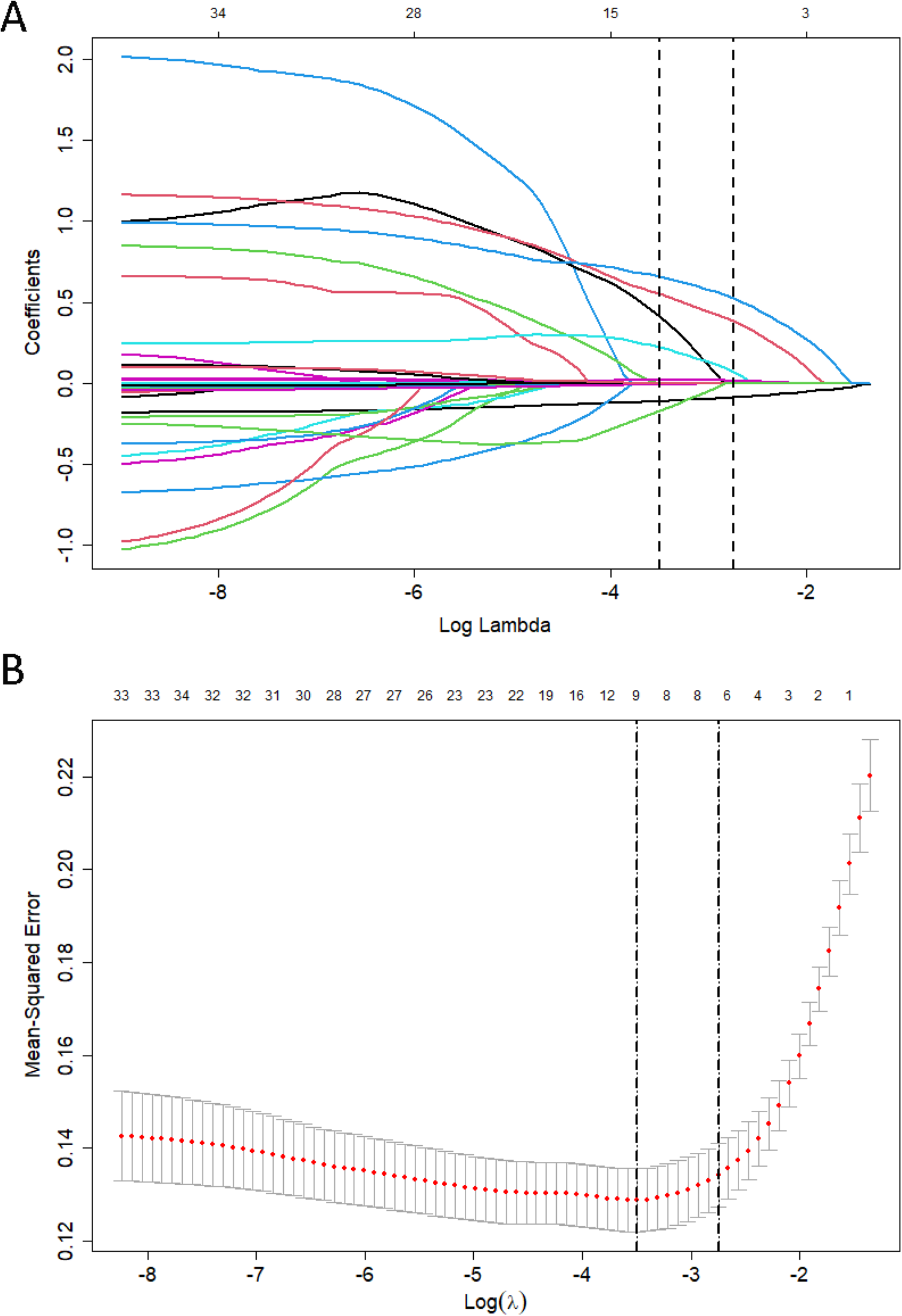
LASSO regression to select feature variables. (A), The curve of the regression coefficient varies with the change of Log(λ) (B) LASSO’s mean squared error changes with Log(λ) in the regression analysis graph. Nine variables with nonzero coefficients were selected by optimal lambda.

**Figure 4.**
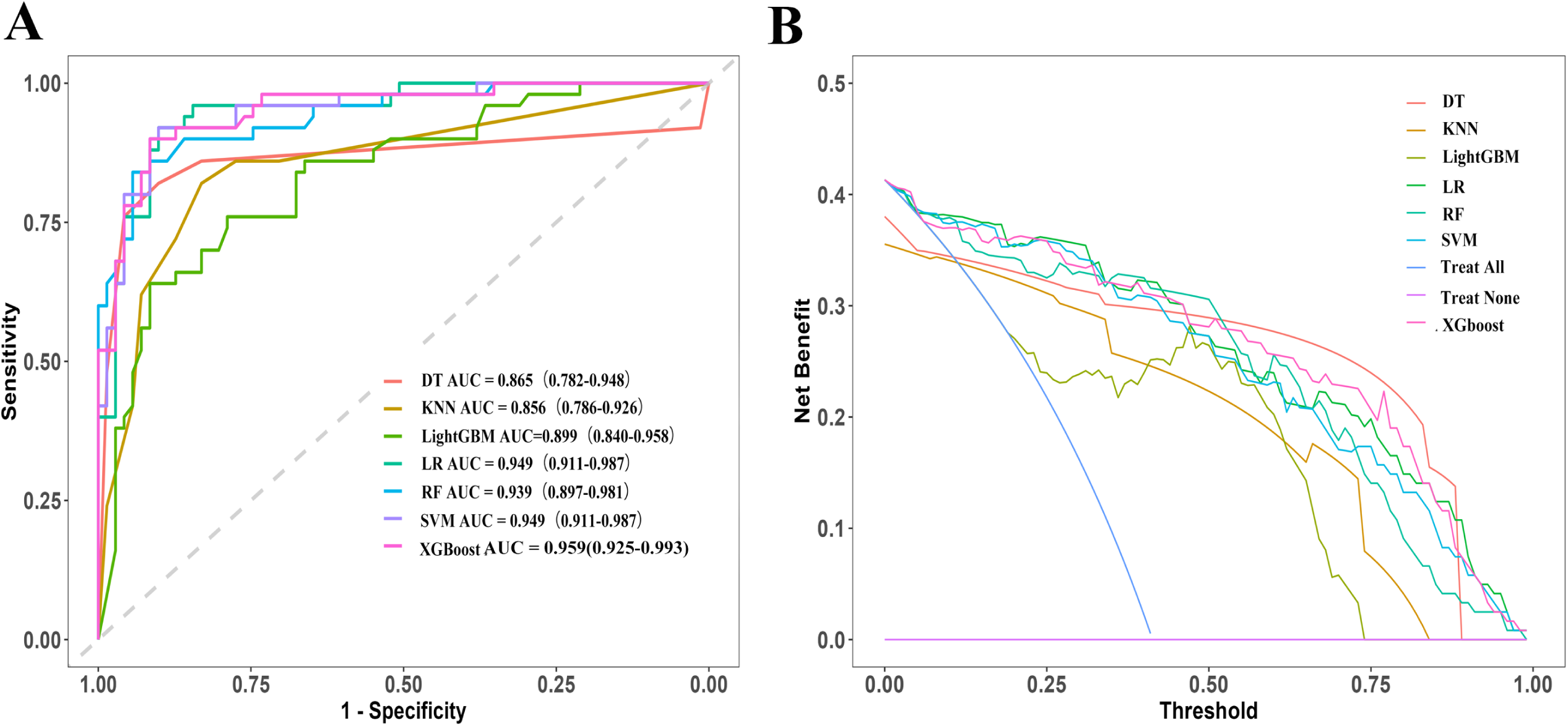
(A)The diagnostic accuracy of machine learning models for detecting left atrial thrombus/spontaneous echo contrast in validation cohorts. (B) The decision curve analysis of the machine learning model demonstrated the net benefits in predicting left atrial thrombus or spontaneous echo contrast.

In the evaluation of LAT/SEC prediction models, ten-fold cross-validation was employed to assess the performance of various ML models. The assessment involved calculating key metrics for each model, including accuracy, precision, recall, F1 score, and AUC values, presented in Table 2. Additionally, ROC curves were plotted for these ML models based on their performance on the test set.

**Table 2.** The diagnostic efficacy of machine learning models in detecting left atrial thrombus or spontaneous echo contrast.

| Model | Accuracy | Precision | Recall | F1 | AUC ( 95% CI ) |
| --- | --- | --- | --- | --- | --- |
| LR | 0.893 | 0.836 | 0.920 | 0.876 | 0.949 ( 0.911-0.987 ) |
| DT | 0.876 | 0.760 | 0.927 | 0.835 | 0.865 ( 0.782-0.948 ) |
| RF | 0.893 | 0.911 | 0.820 | 0.863 | 0.939 ( 0.897-0.981 ) |
| XGBoost | 0.884 | 0.891 | 0.820 | 0.854 | 0.959 ( 0.925-0.993 ) |
| SVM | 0.901 | 0.880 | 0.880 | 0.880 | 0.949 ( 0.911-0.987 ) |
| KNN | 0.810 | 0.800 | 0.720 | 0.758 | 0.856 ( 0.786-0.926 ) |
| LightGBM | 0.860 | 0.851 | 0.800 | 0.825 | 0.899 ( 0.840-0.958 ) |

DT: Decision Tree, KNN: K-Nearest Neighbors, LightGBM: Light Gradient Boosting Machine, LR: Logistic Regression, RF: Random Forest, SVM: Support Vector Machine, XGBoost: eXtreme Gradient Boosting,

From the comparative analysis of the model evaluation results and ROC curves, it was observed that among the seven models assessed, XGBoost demonstrated superior AUC values of 0.959 (95% CI 0.925–0.993) than other ML models. Compared with LR (AUC 0.949, 95% CI 0.911–0.987), the XGBoost model exhibited a slightly more accurate prediction for LAT/SEC risk in NVAF patients (*P* < 0.05). The XGBoost model outperformed the others, presenting the highest clinical net benefit within the threshold probability range of 0.1 to 1.0. The results were shown in Figure 3.

The internal logic of the XGBoost model was analyzed to determine the ranking of the importance of feature variables. It identified PALS, left atrial acceleration factor (α), and 3D-SI as the most critical risk factors for thrombus formation. Other risk factors were ranked in descending order of importance and EF, LAVI, pulmonary vein S/D ratio, prior stroke/TIA, and persistent AF. These findings were visually presented in Figure 5A.

**Figure 5.**
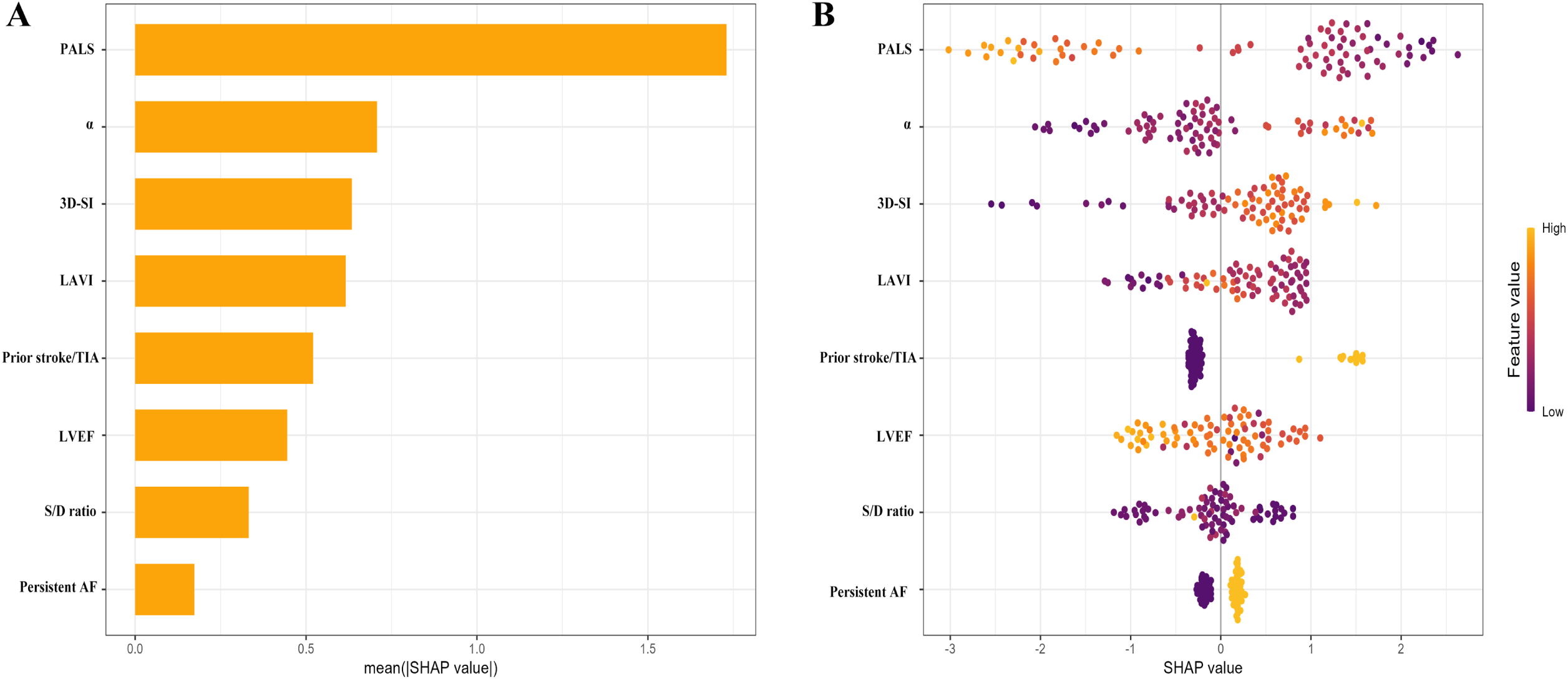
(A) Feature importance view based on XGBoost model. (B) Summary chart of SHAP based on XGBoost model

This study employed the SHAP methodology to perform a detailed analysis of the thrombosis risk prediction model for patients with NVAF, which was developed using the XGBoost algorithm. Figure 5B illustrated a summary plot that conveyed the average influence of each feature on the model’s predictions. In this plot, features represented in the yellow region are those that tend to increase the probability of thrombosis in NVAF patients. For instance, a higher left atrial volume index or the presence of persistent AF were associated with a heightened risk of stroke. Conversely, features in the red region suggested a protective effect against thrombosis. An example of this was the LVEF and PALS, which are negatively correlated with the risk of LAT/SEC.

Beyond a global interpretation, the SHAP values were capable of providing insights into individual risk assessments, as depicted in Figure 6 which shows the factors influencing the thrombosis risk classification for a single patient within the XGBoost model. This individualized analysis not only demonstrated the role each attribute played in predicting thrombosis risk but also indicated its specific impact, aiding clinical practitioners in evaluating the thrombosis risk for particular patients. The significance of each feature was denoted by the magnitude of the arrows, with yellow arrows indicating features that amplify risk and red arrows denoting those that mitigate it.

**Figure 6.**
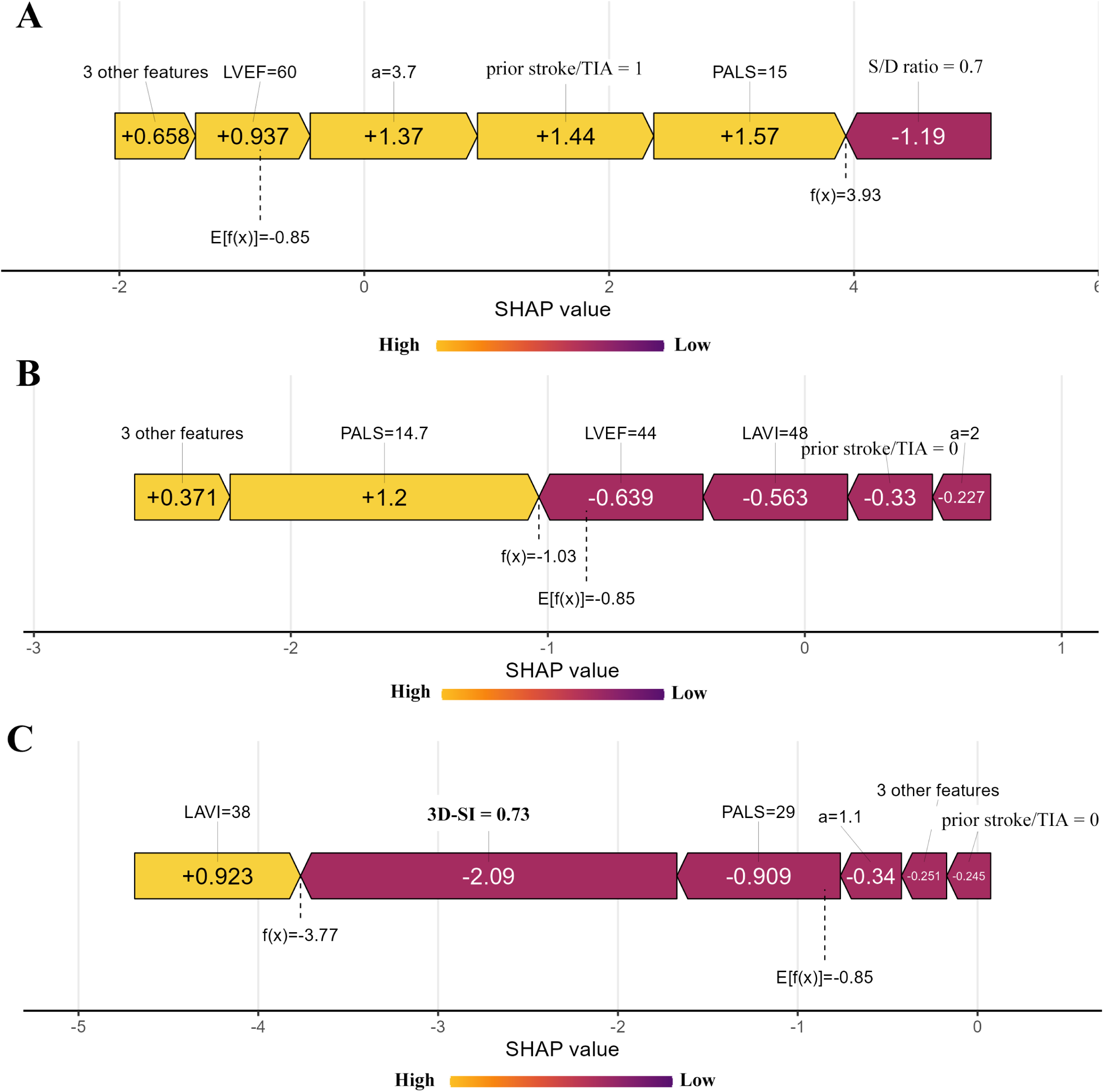
XGBoost model predicting the risk score of thrombosis, with example SHAP charts for certain cases: (A) Example of high-risk thrombosis, (B) and (C) examples of low-risk thrombosis.

For illustration, consider a patient highlighted in Figure 6A with an f(x) value of 3.93, which fallen above the established risk threshold of −0.85, marking the individual as high risk for thrombosis. This patient had a mix of risk-increasing (yellow) and risk-decreasing (red) factors, whose combined effect dictates the overall risk evaluation. The S/D ratio inhibited the risk of thrombosis, while PLAS, LVEF, LAVI, 3D-SI, pre-stroke/TIA, and persistent AF promoted the risk. The comprehensive and quantitative analysis of each factor’s contribution allowed for individualized and precise treatment decisions to be formulated.

## Discussion

This study explored the effectiveness of seven different ML models in predicting the risk of thrombosis in patients with NVAF based on multimodal ultrasound parameters. The results showed that the XGBoost model slightly outperformed the traditional Logistic regression model in predicting the risk of thrombosis in NVAF patients, and demonstrated superior predictive ability compared to other ML algorithms. Left atrial structure (LAVI, 3D-SI), hemodynamic parameters (left atrial acceleration factor and S/D ratio) and functional parameters (PALS, LVEF) were identified as important features for predicting the risk of thrombosis in NVAF patients. Decreased PALS is the most important risk factor for predicting thrombosis. Utilizing multimodal echocardiographic parameters combined with clinical risk factors such as persistent AF and prior stroke/TIA can significantly improve the predictive ability of thrombosis risk in NVAF patients.

Thrombus formation in the left atrium/left atrial appendage is the fundamental cause of stroke in NVAF patients.^18^ Assessing the thrombosis risk in NVAF patients is a crucial step for preventing stroke and improving patient outcomes. Although the CHA_2_DS_2_-VASc score provides good evaluation of stroke risk based on common clinical risk factors in daily practice, but stroke still occurred in patients with a CHA_2_DS_2_-VASc score less than 1.^19^ Previous guidelines do not routinely recommend antithrombotic therapy for patients with a CHA_2_DS_2_-VASc score less than one. Therefore, it is crucial in clinical practice to identify NVAF patients with low CHA_2_DS_2_-VASc scores but significant thrombosis risk.^3^

Combining clinical risk factors with non-invasive echocardiographic parameters may help identify NVAF patients with higher thrombotic risk.^5,20^ Numerous studies have shown that echocardiographic evaluation of left atrial volume and function in NVAF patients can effectively predict the risk of stroke.^21,22^ Previous studies have indicated that left atrial size does not have predictive value for stroke when adjusted for clinical risk factors.^7^ Therefore, solely relying on left atrial diameter may not sensitively predict the risk of thrombus in NVAF patients.^23^ It is recommended by guidelines to integrate advanced echocardiographic parameters into the CHA_2_DS_2_-VASc scores in order to improve the risk stratification of thrombus in patients with NVAF.

Left atrial strain analysis has the capability to detect nuanced alterations in left atrial structure and function that may not be easily discernible through conventional ECG or other cardiac morphological assessments.^24^ Left atrial enlargement typically occurs in later stages of left atrial dysfunction, while left atrial strain measurements can detect early-stage left atrial dysfunction, especially when left atrial volume is still within the normal range, and provide additional prognostic information for cardiovascular disease.^25^ Prior studies have suggested that left atrial strain parameters may be better predictors of adverse cardiovascular events in NVAF patients. ^26,27^ Left atrial strain analysis can help improve risk stratification for NVAF and guide secondary prevention strategies, independent of conventional echocardiographic parameters such as left atrial volume.^21,28^ Using left atrial strain may improve risk stratification and decision-making in patients with cardio embolism, particularly in long-term rhythm monitoring and/or empirical anticoagulant therapy.^29^ Our research also indicated that PALS assessed by speckle tracking echocardiography can identify left atrial dysfunction, closely related to thrombosis risk and independent of other cardiovascular risk factors. AF can cause hemodynamic changes in the atrium, increasing the risk of stroke and other thromboembolic events. The irregular contractions during AF impair the atrium’s pumping function, leading to decreased blood flow speed and increased risk of blood stasis and thrombus formation in patients with NVAF^30^. Prior research has demonstrated a strong link between LAT/SEC and left atrial hemodynamic parameters^31^. Fang et al. discovered that these parameters can help assess stroke risk in NVAF patients by indicating the association between thrombosis risk and blood flow status in the left atrium and left atrial appendage^18^. This provides a foundation for more precise evaluation of stroke risk in NVAF patients. Our study found that left atrial acceleration factor(α) and pulmonary vein S/D ratio were independently correlated with the risk of thrombosis. The previous study also suggested that left atrial sphericity was linked to a prior history of stroke in patients with AF.^32^ Our study also indicated that 3D-SI can predict thrombus risk by showing left atrium morphological changes, regardless of size and without relying on geometric assumptions. A detailed assessment of left atrial function using echocardiography has been shown to be beneficial in assessing risk of thrombosis. This study suggested that left atrial structure (LAVI, 3D-SI), hemodynamic parameters (left atrial acceleration factor a), and functional parameters (PALS, LVEF) were important features for predicting the risk of thrombus in NVAF patients, with decreased PALS being the most significant risk factor for predicting thrombosis.

ML has great potential for innovation and disruption in the medical field. It has been successfully applied in various cardiovascular diseases and has shown promising results in predicting efficacy through visual evaluation.^33,34^ One important application area is managing the potential thrombotic risk in NVAF.^8^ Novel technologies like ML have the potential to significantly greatly enhance stroke risk management and clinical prognosis in patients by identifying those at high risk of thrombosis and intervening early. Previous studies have shown that ML models can accurately predict left atrial appendage thrombosis and outperform conventional stroke risk scores, which may help predict the risk of stroke in NVAF patients.^11^ The internal logic of ML models can identify critical risk factors such as left atrial and left atrial appendage structures and certain biomarkers, which are crucial for predicting thrombosis and optimizing treatment strategies for NVAF patients, with significant implications for preventing ischemic stroke. Studies have used data from the Korean AF Registry of Ischemic Stroke Patients and the Korean University Stroke Registry to internally and externally validate ML models. The results showed that ML models performed well in predicting both outcomes. For adverse outcomes at 3 months, the multilayer perceptron model demonstrated significantly higher ROC values of 0.890 and 0.859 in the internal and external validation groups, respectively, outperforming logistic regression^35^. Therefore, explainable ML models can effectively predict short-term outcomes and identify high-risk NVAF-related stroke patients.

XGBoost is an advanced decision tree ensemble technique widely used in the medical field. It is highly regarded for its excellent classification performance, ability to model complex nonlinear relationships, and handle high-dimensional data.^36^ In the process of building predictive models, model interpretability is also an important factor. The XGBoost model introduces tree-based model feature importance and more complex SHAP values to reveal the specific contribution of each feature to the prediction results, balancing model performance and interpretability. This study utilized the internal logic of the XGBoost model to establish a predictive model for thrombosis risk in NVAF patients, demonstrating good performance in model evaluation. In the validation set, the AUC value for predicting LAT/SEC was 0.959 (95% CI 0.959-0.993), outperforming other ML models. Feature importance analysis and SHAP interpretability analysis were used to rank the features based on their importance and frequency in each model, identifying the risk factors for thrombus in NVAF. Through interpreting SHAP plots with actual cases, clinical physicians can better understand how the model operates and guide personalized treatment strategies for NVAF patients.

In this research, the utilization of advanced machine learning algorithms to combine multimodal echocardiographic parameters with clinical risk factors resulted in a substantial improvement in the predictive accuracy for evaluating thrombus risk in patients with NVAF. Besides, XGBoost model demonstrated superior predictive performance, aiding in the precise clinical assessment of thrombus risk in NVAF patients. The model showed that decreased PALS, hemodynamic abnormalities and left atrium spherical remodeling were significant factors correlated with increased risk of thrombus in NVAF.

## Limitations

This study had a relatively small sample size and lacked external validation, requiring an increase in sample size for multicenter verification to enhance the stability and generalizability of the model. In this study, only a subset of clinical and echocardiographic features was considered, without incorporating more detailed biochemical indicators, possibly overlooking other potential risk factors. ML are designed to uncover the latent connections between input variables and output values; however, they may not be able to capture relationships that are not fully elucidated due to specific variables. Additionally, anticoagulants offer notable benefits in the prevention of left atrial appendage thrombus. However, in this study, only the type of anticoagulant medication was considered, without taking into account dosage and duration of irregular medication records. Finally, this study included more SEC patients and fewer patients with LAT, leading to some selection bias and potential deviations in predictive results. Additionally, there may be differences in thrombus formation risk among SEC patients of different levels.

## Conclusion

This study demonstrates that the integration of multimodal echocardiographic parameters with clinical risk factors based on sophisticated ML algorithms significantly improved the predictive accuracy of thrombus risk in patients with NVAF. Specifically, the XGBoost model slightly outperforms the traditional Logistic regression model in predicting thrombus formation risk in NVAF patients, and shows superior predictive ability compared to other ML algorithms. Left atrial structure (LAVI, 3D-SI), hemodynamic indices (left atrial acceleration factor and S/D ratio) and functional parameters (PALS, LVEF) are crucial factors in predicting the risk of thrombus formation in patients with non-valvular atrial fibrillation (NVAF). Among these factors, PALS emerges as the most important risk factor for predicting LAT/SEC. Developing a predictive model utilizing machine learning techniques that incorporate multimodal echocardiographic parameters in conjunction with clinical risk factors, such as persistent atrial fibrillation and prior stroke/TIA, has the potential to enhance the predictive accuracy of thrombosis risk in individuals with NVAF.

## Data Availability

All pertinent data for the study have been included in the article and are available from the corresponding author upon reasonable request.

## Founding

The research was funded by the Key Program of Guangxi Natural Science Foundation (Grant No. 2023GXNSFDA026010), Youth Science Foundation of Guangxi Medical University (Grant No. GXMUYSF201916), and the ‘139’ Project of Guangxi Medical aimed at training high-level backbone talents (Grant No. G201903014).

## Reference

1. Joglar J, Chung M, Armbruster A, Benjamin E, Chyou J, Cronin E, Deswal A, Eckhardt L, Goldberger Z, Gopinathannair R, et al. 2023 ACC/AHA/ACCP/HRS Guideline for the Diagnosis and Management of Atrial Fibrillation: A Report of the American College of Cardiology/American Heart Association Joint Committee on Clinical Practice Guidelines. Circulation. 2024;149:e1–e156. doi: 10.1161/cir.0000000000001193

2. Migdady I, Russman A, Buletko A. Atrial Fibrillation and Ischemic Stroke: A Clinical Review. Seminars in neurology. 2021;41:348–364. doi: 10.1055/s-0041-1726332

3. Jagadish P, Kabra R. Stroke Risk in Atrial Fibrillation: Beyond the CHADS-VASc Score. Current cardiology reports. 2019;21:95. doi: 10.1007/s11886-019-1189-6

4. Hijazi Z, Oldgren J, Lindbäck J, Alexander J, Connolly S, Eikelboom J, Ezekowitz M, Held C, Hylek E, Lopes R, et al. The novel biomarker-based ABC (age, biomarkers, clinical history)-bleeding risk score for patients with atrial fibrillation: a derivation and validation study. *Lancet (London*, England). 2016;387:2302–2311. doi: 10.1016/s0140-6736(16)00741-8

5. Lee J, Cha M, Nam G, Choi K, Sun B, Kim D, Song J, Kang D, Song J, Cho M. Incidence and predictors of left atrial thrombus in patients with atrial fibrillation under anticoagulation therapy. Clinical research in cardiology : official journal of the German Cardiac Society. 2024. doi: 10.1007/s00392-024-02422-5

6. Chen J, Zhou M, Wang H, Zheng Z, Rong W, He B, Zhao L. Risk factors for left atrial thrombus or spontaneous echo contrast in non-valvular atrial fibrillation patients with low CHADS-VASc score. Journal of thrombosis and thrombolysis. 2022;53:523–531. doi: 10.1007/s11239-021-02554-9

7. Leung M, van Rosendael P, Abou R, Ajmone Marsan N, Leung D, Delgado V, Bax J. Left atrial function to identify patients with atrial fibrillation at high risk of stroke: new insights from a large registry. European heart journal. 2018;39:1416–1425. doi: 10.1093/eurheartj/ehx736

8. Wegner FK, Plagwitz L, Doldi F, Ellermann C, Willy K, Wolfes J, Sandmann S, Varghese J, Eckardt L. Machine learning in the detection and management of atrial fibrillation. Clin Res Cardiol. 2022;111:1010–1017. doi: 10.1007/s00392-022-02012-3

9. undefined u, undefined u, undefined u, undefined u, undefined u, undefined u, undefined u, undefined u, undefined u, undefined u, et al. Artificial intelligence in the risk prediction models of cardiovascular disease and development of an independent validation screening tool: a systematic review. BMC Med. 2024;22. doi: 10.1186/s12916-024-03273-7

10. Chahine Y, Magoon MJ, Maidu B, Del Álamo JC, Boyle PM, Akoum N. Machine Learning and the Conundrum of Stroke Risk Prediction. Arrhythm Electrophysiol Rev. 2023;12:e07. doi: 10.15420/aer.2022.34

11. Zhao Y, Cao LY, Zhao YX, Wang F, Xie LL, Xing HY, Wang Q. Medical record data-enabled machine learning can enhance prediction of left atrial appendage thrombosis in nonvalvular atrial fibrillation. Thromb Res. 2023;223:174–183. doi: 10.1016/j.thromres.2023.01.001

12. Hundemer GL, White CA, Norman PA, Knoll GA, Tangri N, Sood MM, Hiremath S, Burns KD, McCudden C, Akbari A. Performance of the 2021 Race-Free CKD-EPI Creatinine- and Cystatin C-Based Estimated GFR Equations Among Kidney Transplant Recipients. Am J Kidney Dis. 2022;80:462–472.e461. doi: 10.1053/j.ajkd.2022.03.014

13. Lang R, Badano L, Mor-Avi V, Afilalo J, Armstrong A, Ernande L, Flachskampf F, Foster E, Goldstein S, Kuznetsova T, et al. Recommendations for cardiac chamber quantification by echocardiography in adults: an update from the American Society of Echocardiography and the European Association of Cardiovascular Imaging. Journal of the American Society of Echocardiography : official publication of the American Society of Echocardiography. 2015;28:1–39.e14. doi: 10.1016/j.echo.2014.10.003

14. Reiter G, Kovacs G, Reiter C, Schmidt A, Fuchsjäger M, Olschewski H, Reiter U. Left atrial acceleration factor as a magnetic resonance 4D flow measure of mean pulmonary artery wedge pressure in pulmonary hypertension. Frontiers in cardiovascular medicine. 2022;9:972142. doi: 10.3389/fcvm.2022.972142

15. undefined u, undefined u, undefined u, undefined u, undefined u, undefined u, undefined u, undefined u, undefined u, undefined u, et al. Guidelines for Developing and Reporting Machine Learning Predictive Models in Biomedical Research: A Multidisciplinary View. J Med Internet Res. 2016;18. doi: 10.2196/jmir.5870

16. undefined u, undefined u, undefined u, undefined u, undefined u, undefined u, undefined u, undefined u, undefined u, undefined u, et al. Protocol for development of a reporting guideline (TRIPOD-AI) and risk of bias tool (PROBAST-AI) for diagnostic and prognostic prediction model studies based on artificial intelligence. BMJ Open. 2021;11. doi: 10.1136/bmjopen-2020-048008

17. undefined u, undefined u, undefined u, undefined u, undefined u, undefined u, undefined u, undefined u, undefined u. Comparative performance analysis of Boruta, SHAP, and Borutashap for disease diagnosis: A study with multiple machine learning algorithms. Network. 2024. doi: 10.1080/0954898x.2024.2331506

18. Fang R, Wang Z, Zhao X, Wang J, Li Y, Zhang Y, Chen Q, Wang J, Liu Q, Chen M, et al. Stroke risk evaluation for patients with atrial fibrillation: Insights from left atrial appendage with fluid-structure interaction analysis. Computers in biology and medicine. 2022;148:105897. doi: 10.1016/j.compbiomed.2022.105897

19. Kim T, Yang P, Kim D, Yu H, Uhm J, Kim J, Pak H, Lee M, Joung B, Lip G. CHADS-VASc Score for Identifying Truly Low-Risk Atrial Fibrillation for Stroke: A Korean Nationwide Cohort Study. Stroke. 2017;48:2984–2990. doi: 10.1161/strokeaha.117.018551

20. Segan L, Nanayakkara S, Spear E, Shirwaiker A, Chieng D, Prabhu S, Sugumar H, Ling L, Kaye D, Kalman J, et al. Identifying Patients at High Risk of Left Atrial Appendage Thrombus Before Cardioversion: The CLOTS-AF Score. Journal of the American Heart Association. 2023;12:e029259. doi: 10.1161/jaha.122.029259

21. Leong D, Joyce E, Debonnaire P, Katsanos S, Holman E, Schalij M, Bax J, Delgado V, Marsan N. Left Atrial Dysfunction in the Pathogenesis of Cryptogenic Stroke: Novel Insights from Speckle-Tracking Echocardiography. Journal of the American Society of Echocardiography : official publication of the American Society of Echocardiography. 2017;30:71–79.e71. doi: 10.1016/j.echo.2016.09.013

22. Lin W, Xue Y, Liu F, Fang X, Zhan X, Liao H, Tse G, Wu S. Left atrial enlargement and non-paroxysmal atrial fibrillation as risk factors for left atrial thrombus/spontaneous Echo contrast in patients with atrial fibrillation and low CHADS-VASc score. Journal of geriatric cardiology : JGC. 2020;17:155–159. doi: 10.11909/j.issn.1671-5411.2020.03.001

23. Bisbal F, Gómez-Pulido F, Cabanas-Grandío P, Akoum N, Calvo M, Andreu D, Prat-González S, Perea R, Villuendas R, Berruezo A, et al. Left Atrial Geometry Improves Risk Prediction of Thromboembolic Events in Patients With Atrial Fibrillation. Journal of cardiovascular electrophysiology. 2016;27:804–810. doi: 10.1111/jce.12978

24. Aga Y, Abou Kamar S, Chin J, van den Berg V, Strachinaru M, Bowen D, Frowijn R, Akkerhuis M, Constantinescu A, Umans V, et al. Potential role of left atrial strain in estimation of left atrial pressure in patients with chronic heart failure. ESC heart failure. 2023;10:2345–2353. doi: 10.1002/ehf2.14372

25. Mukai Y, Nakanishi K, Daimon M, Iwama K, Yoshida Y, Hirose K, Yamamoto Y, Seki H, Nakao T, Oshima T, et al. Prevalence, Associated Factors, and Echocardiographic Estimation of Left Atrial Hypertension in Patients With Atrial Fibrillation. Journal of the American Heart Association. 2023;12:e030325. doi: 10.1161/jaha.123.030325

26. Bashir Z, Chen E, Wang S, Shu L, Goldstein E, Rana M, Kala N, Dai X, Mandel D, Has P, et al. Left atrial strain, embolic stroke of undetermined source, and atrial fibrillation detection. Echocardiography (Mount Kisco, NY). 2024;41:e15738. doi: 10.1111/echo.15738

27. Mannina C, Ito K, Jin Z, Yoshida Y, Matsumoto K, Shames S, Russo C, Elkind M, Rundek T, Yoshita M, et al. Association of Left Atrial Strain With Ischemic Stroke Risk in Older Adults. JAMA cardiology. 2023;8:317–325. doi: 10.1001/jamacardio.2022.5449

28. Hauser R, Nielsen A, Skaarup K, Lassen M, Duus L, Johansen N, Sengeløv M, Marott J, Jensen G, Schnohr P, et al. Left atrial strain predicts incident atrial fibrillation in the general population: the Copenhagen City Heart Study. European heart journal Cardiovascular Imaging. 2021;23:52–60. doi: 10.1093/ehjci/jeab202

29. Vera A, Cecconi A, Ximénez-Carrillo Á, Ramos C, Martínez-Vives P, Lopez-Melgar B, Sanz-García A, Ortega G, Aguirre C, Montes Á, et al. Left Atrial Strain Predicts Stroke Recurrence and Death in Patients With Cryptogenic Stroke. Am J Cardiol. 2024;210:51–57. doi: 10.1016/j.amjcard.2023.10.001

30. Doukky R, Garcia-Sayan E, Patel M, Pant R, Wassouf M, Shah S, D’Silva O, Kehoe R. Impact of Diastolic Function Parameters on the Risk for Left Atrial Appendage Thrombus in Patients with Nonvalvular Atrial Fibrillation: A Prospective Study. Journal of the American Society of Echocardiography : official publication of the American Society of Echocardiography. 2016;29:545–553. doi: 10.1016/j.echo.2016.01.014

31. Stojadinović P, Deshraju A, Wichterle D, Fukunaga M, Peichl P, Kautzner J, Šramko M. The hemodynamic effect of simulated atrial fibrillation on left ventricular function. Journal of cardiovascular electrophysiology. 2022;33:2569–2577. doi: 10.1111/jce.15669

32. Dudzińska-Szczerba K, Zalewska M, Niemiro W, Michałowska I, Piotrowski R, Sikorska A, Kułakowski P, Baran J. Association of Left Atrial Sphericity with Risk of Stroke in Patients with Atrial Fibrillation. Sub-Analysis of the ASSAM Study. Cardiovasc Eng Technol. 2022;13:419–427. doi: 10.1007/s13239-021-00587-y

33. Raghunath S, Pfeifer JM, Ulloa-Cerna AE, Nemani A, Carbonati T, Jing L, vanMaanen DP, Hartzel DN, Ruhl JA, Lagerman BF, et al. Deep Neural Networks Can Predict New-Onset Atrial Fibrillation From the 12-Lead ECG and Help Identify Those at Risk of Atrial Fibrillation-Related Stroke. Circulation. 2021;143:1287–1298. doi: 10.1161/circulationaha.120.047829

34. Bifulco SF, Macheret F, Scott GD, Akoum N, Boyle PM. Explainable Machine Learning to Predict Anchored Reentry Substrate Created by Persistent Atrial Fibrillation Ablation in Computational Models. J Am Heart Assoc. 2023;12:e030500. doi: 10.1161/jaha.123.030500

35. Jeon E, Jung S, Yeo T, Seo W, Jung J. Predicting short-term outcomes in atrial-fibrillation-related stroke using machine learning. Frontiers in neurology. 2023;14:1243700. doi: 10.3389/fneur.2023.1243700

36. Zou Y, Shi Y, Sun F, Liu J, Guo Y, Zhang H, Lu X, Gong Y, Xia S. Extreme gradient boosting model to assess risk of central cervical lymph node metastasis in patients with papillary thyroid carcinoma: Individual prediction using SHapley Additive exPlanations. Computer methods and programs in biomedicine. 2022;225:107038. doi: 10.1016/j.cmpb.2022.107038

